# Multi-Omics Causal Inference Identifies *Phocaeicola vulgatus*–Mediated 5’-Methylthioadenosine Clearance Contributing to Mitochondrial Protection in Heart Failure

**DOI:** 10.1101/2025.10.23.25338456

**Authors:** Sheng Gao, Min Yang, Yingyuan Lu, Xiaoqian Sun, Yixuan Qian, Huijuan Wang, Tingfang Chen, Zhi Yang, Zhiyong Du, Yulin Ouyang, Yong Wang, Wei Wang, Chun Li

## Abstract

**Background:** Heart failure (HF) is a multifactorial metabolic disorder. While gut microbial dysbiosis is increasingly implicated in HF, the specific causal microbes and their molecular mediators linking intestinal perturbations to cardiac dysfunction remain undefined.

**Methods:** We performed integrated fecal 16S rRNA and serum metabolomic profiling in a clinical HF cohort comprising 149 HF patients and 50 healthy controls. We systematically assessed the multidimensional alterations and diagnostic potential of the gut microbiome and serum metabolome. Key microbe-metabolite interactions were identified through causal inference and experimentally validated using i*n vitro* bacterial cultures, *in vivo* mouse models of HF, and assays of mitochondrial function.

**Results:** HF patients exhibited significant alterations in the gut microbiome and serum metabolome related to energy homeostasis, vascular tone regulation, and inflammatory balance. These differential microbial and metabolic signatures demonstrated superior diagnostic potential for HF. A key finding was depletion of the commensal bacterium *Phocaeicola vulgatus* led to the accumulation of serum 5’-methylthioadenosine (MTA), which in turn induced mitochondrial dysfunction and aggravated cardiac injury. Restoring *P. vulgatus* mitigated these effects by metabolizing MTA, whereas direct MTA administration recapitulated mitochondrial dysfunction and exacerbated HF pathology.

**Conclusions:** This study provides an integrative multi-omics perspective on the gut microbiome-serum metabolome interplay in HF, revealing both diagnostic biomarkers and mechanistic insights. Through a causal inference framework, we identify the *P. vulgatus*–MTA axis as a causal pathway through which gut microbes influence HF progression.

## Introduction

Heart failure (HF) is a complex clinical syndrome that represents the final stage of structural or functional cardiac impairment, characterized by profound metabolic reprogramming, systemic inflammation, and multi-organ dysfunction ^[1, 2^]. Despite advances in pharmacological and device-based therapies, HF remains a major global health burden with persistently high morbidity and mortality ^[3, 4^]. Accumulating evidence indicates that the gut microbiota acts as an environmental modifier of cardiovascular health, forming a bidirectional “gut–heart axis” that modulates cardiac function through metabolic and inflammatory signaling ^[5–7]^. However, the specific causal microbial taxa and effector metabolites that link gut perturbations to cardiac dysfunction remain largely undefined.

Microbiota-derived metabolites serve as key mediators of gut–host communication in cardiovascular disease ^[7–9]^. For instance, trimethylamine N-oxide (TMAO) promotes myocardial inflammation and mitochondrial injury ^[8]^, whereas depletion of short-chain fatty acid–producing genera such as *Lachnospiraceae* and *Ruminococcaceae* impairs intestinal barrier function and exacerbates HF ^[9]^. Notably, HF-associated microbial signatures have been observed in metabolically dysregulated individuals prior to disease onset, implying a potential causal role of microbial metabolism in early HF development ^[10]^. Therefore, future efforts are needed to characterize microbial signatures and their contributions to the circulating metabolism dysbiosis and HF progression.

Causal-based approaches have been widely adopted to elucidate the relationships among the gut microbes, metabolites, and cardiovascular diseases ^[11–14]^, offering a strategy that accounts for potential confounding effects ^[15]^. For example, Huang et al. conducted bidirectional mediation analysis and revealed that the overgrowth of *Enterococcus* contributes to immune-inflammatory imbalance in congenital heart disease via metabolomic perturbations ^[11]^. Similarly, Dai et al. applied mendelian randomization to reveal causal links that *Bacteroides dorei* increases HF risk partly via elevated apolipoprotein B levels ^[12]^. However, these methods rely on predefined assumptions and limited variable scope, reducing their applicability to high-dimensional omics data ^[16]^. In contrast, graph-based causal inference can capture both connectivity and causality from observational data, enabling unbiased inference of interactions in microbe–metabolite networks ^[17, 18^], which is essential for uncovering key microbe–metabolite interactions involved in HF progression.

Here, we integrated gut metagenomic and serum metabolomic data from a clinical HF cohort and applied graph-based causal inference to systematically identify key microbe–metabolite interactions. Through *in vivo* and *in vitro* validation, we revealed a mechanistic link between the commensal bacterium *Phocaeicola vulgatus* and the metabolite 5’-methylthioadenosine (MTA). Depletion of *P. vulgatus* led to serum MTA accumulation, mitochondrial dysfunction, and HF progression, whereas its supplementation restored myocardial mitochondrial and cardiac function. These findings reveal a direct microbe–metabolite–host interaction axis contributing to HF pathogenesis and highlight microbial metabolic modulation as a promising therapeutic strategy.

## Results

### Characteristics of the Chinese multi-omics cohort and study design

We established a clinical cohort of Chinese participants to systematically characterize gut microbiome and serum metabolome alterations in HF. Individuals with gastrointestinal disorders or not meeting the diagnostic criteria for HF were excluded. A total of 199 participants were enrolled, comprising 149 patients with HF and 50 age- and sex-matched healthy controls (HCs) (**Table S1**). Fecal samples underwent 16S rRNA gene sequencing, and paired serum samples were subjected to targeted metabolomic profiling. The overall study design and analytical workflow are summarized in **Figure 1**.

**Figure 1.**
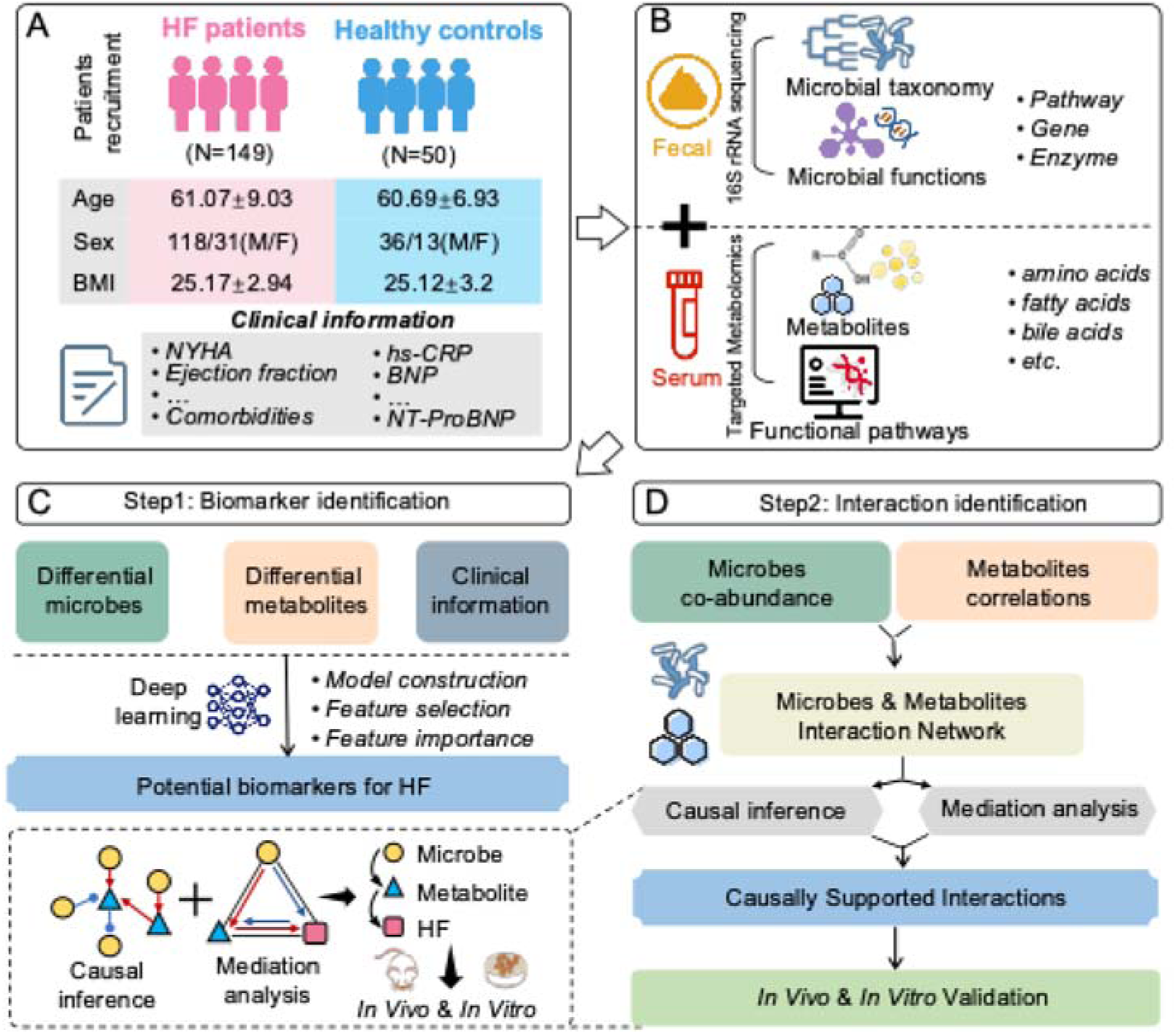
Overview of the study population and analytical workflow. **(A)** A total of 149 HF patients and 50 age- and gender-matched HCs were enrolled. **(B)** Fecal and serum samples were collected for 16S rRNA gene sequencing and targeted metabolomics profiling, respectively. **(C-D)** The main analytical framework of this study included: identification of potential multi-omics biomarkers for HF via a deep learning-based diagnostic model, incorporating feature selection and importance evaluation (C); Construction of a microbe–metabolite interaction network; identification of causal relationships using in silico approaches, including causal inference and mediation analysis; experimental validation of selected interactions through *in vivo* and *in vitro* assays (D).

### Gut microbiome and serum metabolome alterations in HF patients

Potential confounding factors, including age, sex, and BMI, had no significant impact on microbial and metabolic profiles **(Figure S1)** and were further controlled in differential signature analysis.

For gut microbiome, we observed significant differences in both α- and β-diversity, as well as in the overall microbial community composition between HF patients and HCs (**Figure 2A-C, Figure S2)**. Differential signature analysis identified 54 genera with significant changes. Specifically, *Phocaeicola_A_858004*, *Lachnospira*, and *Brevundimonas* were significantly reduced in HF, while *Alistipes_A_871400*, *Blautia_A_141781*, and *Akkermansia* were enriched (**Figure 2D, Table S2)**. At other taxonomic levels, differential signatures are shown in **Figure S3A–D**. Functional analysis revealed 45 microbial pathways upregulated and 40 downregulated in HF, including key metabolic processes such as superpathways of histidine, purine, and pyrimidine biosynthesis, adenosine nucleotide degradation II **(Figure S3E, Table S3)**. At finer resolution, 928 KEGG Orthology (KO) genes and 401 enzymes were differentially abundant **(Figure S3F–G, Tables S4–5)**, largely concordant with pathway-level changes **(Figure S4A–B)**, indicating extensive shifts in microbial metabolic potential associated with HF.

**Figure 2.**
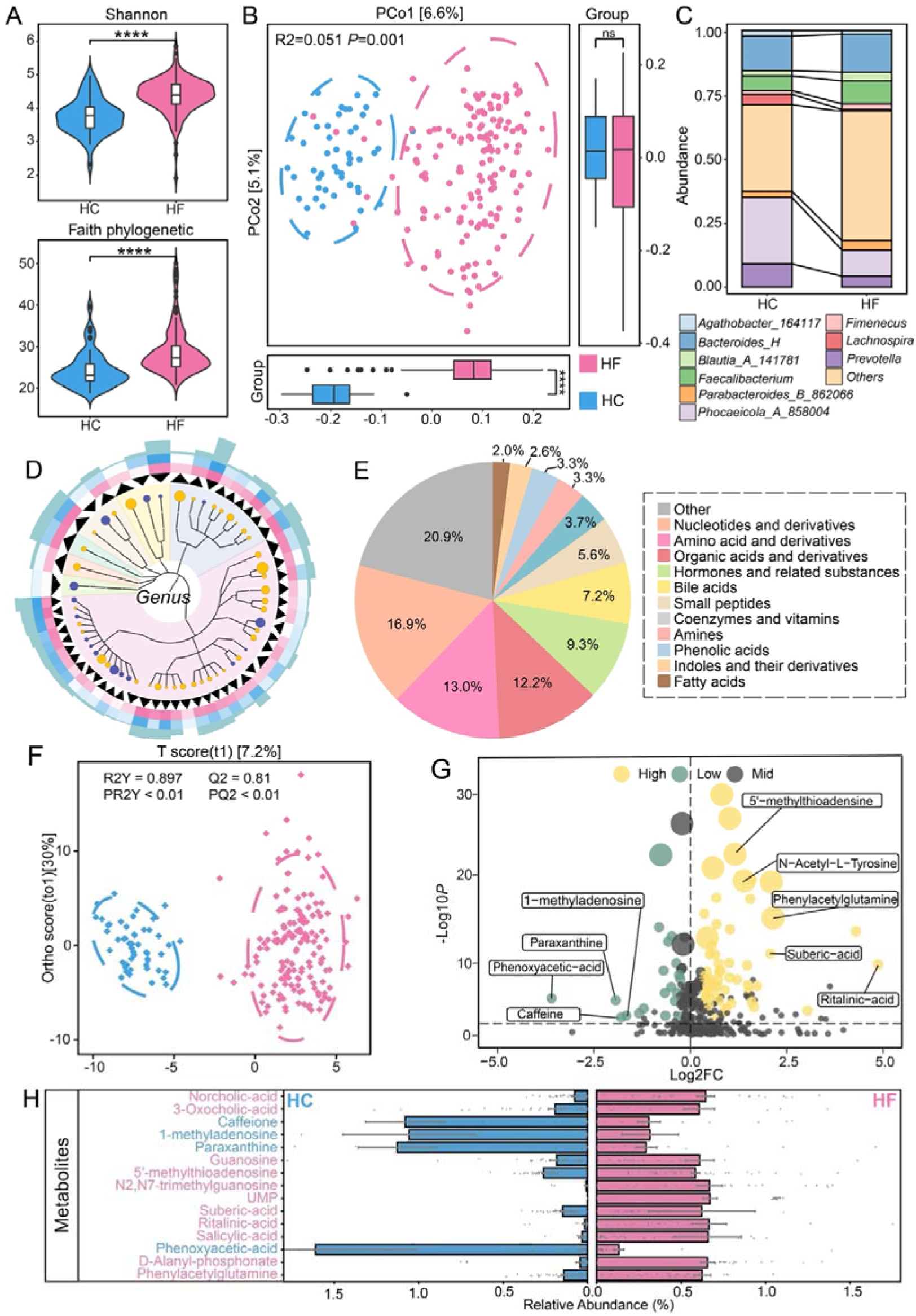
The alterations in the gut microbiome and serum metabolome of HF patients. **(A)** Alpha diversity indices, including Shannon and Faith’s phylogenetic diversity, were calculated for patients with HF (pink, n = 149) and HCs (blue, n = 50). Statistical significance is indicated as follows: *P* < 0.05 (*), *P* < 0.01 (**), *P* < 0.001 (***), and *P* < 0.0001 (****), not significant (ns). **(B)** Principal coordinate analysis (PCoA) based on Bray–Curtis distance revealed distinct microbial community structures between HF patients and HCs (R² = 0.051, *P* = 0.001), as determined by PERMANOVA with 999 permutations (two-sided test). **(C)** The taxonomic composition of the gut microbiome at genus level in HF patients and HCs. **(D)** Differentially abundant genera identified by the Wilcoxon test are visualized on a phylogenetic tree. Node size represents fold-change (HF vs. HC), with orange and purple denoting up- and downregulated taxa, respectively. The innermost ring shows directional arrows (outward = upregulated, inward = downregulated), the middle rings display relative abundances, and the outermost ring indicates LDA scores from LEfSe analysis. **(E)** Pie chart of metabolite classification. Different colors represent distinct metabolite classes, and the percentage indicates the proportion of each class relative to the total number of metabolites. **(F)** The OrthoPLSDA model of serum metabolome analysis was performed on HF patients and HCs (*P*_Q2_< 0.01 and *P*_R2Y_ <0.01). **(G)** The volcano plot of differential metabolites between HF patients and HCs. Yellow, green, and gray dots represent upregulated, downregulated, and non-significant metabolites, respectively. **(H)** The abundances of differential metabolites in HF patients and HCs. Metabolites labeled in pink indicate upregulation in HF, while those in blue indicate downregulation.

Targeted metabolomics quantified 540 metabolites in serum, including 91 nucleotides and derivatives, 70 amino acids and related compounds, 66 organic acids, 50 hormones, 39 bile acids, and additional amines, indoles, and free fatty acids (**Figure 2E)**. Multivariate OPLS-DA analysis identified 83 significantly altered metabolites between HF patients and HCs (63 upregulated, 20 downregulated), including guanosine, MTA, and phenylacetylglutamine (PAGln) (empirical *P-value* for Q² < 0.01 and R²Y < 0.01; **Figure 2F–H, Table S6**). At the functional level, over-representation analysis (ORA) revealed 50 enriched pathways, predominantly related to amino acid metabolism, energy production, purine and pyrimidine metabolism, as well as bile acid and lipid metabolism **(Table S7)**. Notably, many of these metabolic alterations were consistent with microbial functional shifts, highlighting coordinated disruption of energy homeostasis, vascular tone regulation, and inflammatory balance along the gut–host axis.

### Diagnostic potential of gut microbiome and serum metabolome signatures for HF

To evaluate the potential of gut microbiome and serum metabolome for early HF detection, we randomly divided the cohort into training and testing sets (8:2 ratio) and constructed neural network-based classification models using the differential microbial and metabolic signatures. For gut microbiome data, genus-level signatures achieved the best performance (AUC = 0.94), outperforming microbial pathways (AUC = 0.93), KO genes (AUC = 0.86), and enzymes (AUC = 0.85) (**Figure 3A, Figure S5A-E, Table S8)**. Top-ranked genera such as *Lachnospira*, *Blautia_A_141781*, *Phocaeicola_A_858004* contributed the highest SHAP values **(Table S9)**, highlighting their marked alterations in HF and potential involvement in disease pathogenesis. For serum metabolomics data, metabolome-based models achieved even higher predictive accuracy (AUC = 0.96), with top metabolites including D-alanyl-D-alanine, UMP, caffeine, and trans-aconitic acid (**Figure 3A, Figure S5F, Table S10)**. In addition, models based solely on clinical indicators had lower predictive performance (AUC = 0.84), though high-sensitivity troponin I (hsTnI) and B-type natriuretic peptide (BNP) remained the most informative features **(Table S11)**.

**Figure 3.**
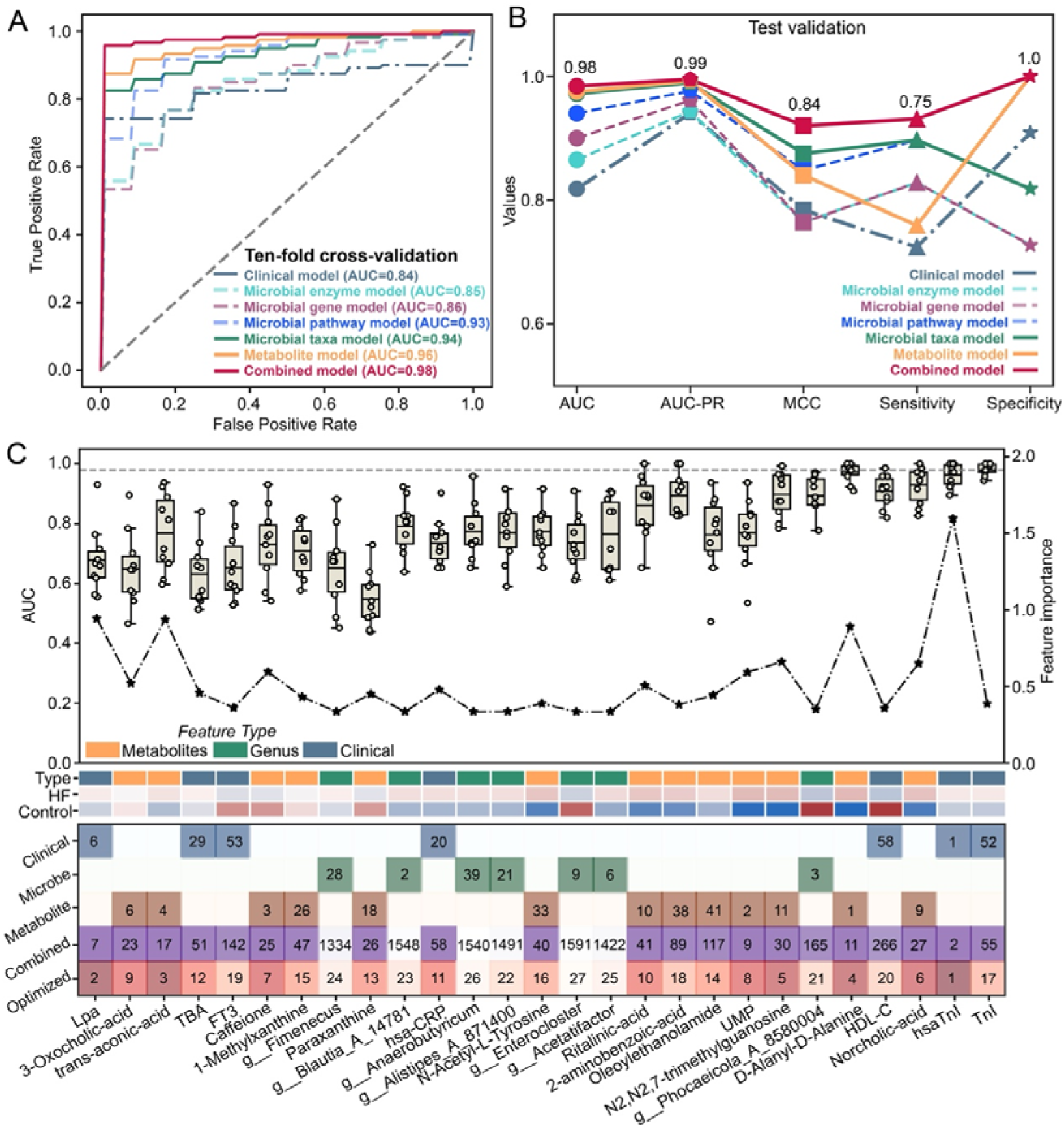
The performance of diagnostic models for HF constructed with multidimensional signatures. **(A)** ROC curves from ten-fold cross-validation of diagnostic models based on clinical, microbial (taxonomy, gene, enzyme), metabolic, and combined signatures. **(B)** Model performance in the validation dataset. **(C)** Recursive feature elimination for the combined model. The boxplot shows cumulative AUC across cross-validation iterations (left y-axis), and the line plot indicates feature importance (right y-axis). Feature categories are color-coded: orange (metabolites), green (microbes), dark blue (clinical). The following two bars denote relative abundance, with red and blue indicating up- and downregulation in HF, respectively. The heatmap below summarizes feature importance rankings across models: columns represent features in the optimized model, and rows represent models built from single data types (e.g., genus, metabolic pathway, serum metabolite, clinical indicator). Darker shading reflects higher importance.

We then constructed an integrative diagnostic model by combining microbial, metabolomic, and clinical features across all levels. The combined model achieved an AUC of 0.98 in both cross-validation and testing validation **(Table S12)**, which underscores the performance gains afforded by multi-omics integration. To enhance clinical utility, we developed an optimized model incorporating only genus-level microbial taxa, serum metabolites, and clinical indicators, which retained comparable accuracy with an average AUC of 0.98 and 0.91 in ten-fold cross-validation and testing dataset, respectively **(Figure S5G-H, Table S13)**. Further, using recursive feature elimination, we identified a minimal feature set of 27 signatures (7 genera, 13 metabolites, 7 clinical indicators), which retained a high predictive performance with an AUC of 0.98 (**Figure 3C)**. Notably, the genus *Phocaeicola_A_858004* ranked among the top microbial contributors, further supporting its strong association with the HF disease state. In sum, these findings suggest that multi-omics signatures, especially those derived from the gut microbiome and serum metabolome, offer superior predictive power and can complement conventional clinical biomarkers for the early identification of individuals at risk of HF.

### Microbe–metabolite causal networks uncover nucleoside metabolism disruption in HF

To investigate gut microbiome–serum metabolome crosstalk, we applied HALLA analysis and identified significant microbe–metabolite associations **(Figure S6A–C)**. *Intestinibacter* and *Akkermansia* showed widespread positive correlations with organic acids and amino acid metabolites, linking them to host energy production and nitrogen metabolism. In particular, *Intestinibacter* was positively associated with L-isoleucine and L-kynurenine, suggesting its role in branched-chain amino acid and tryptophan metabolism—both known to influence cardiovascular function ^[19, 20^]. In contrast, taxa including *Clostridium*, *Enterocloster*, *Ruminococcus_D* (phylum *Firmicutes*), *Phocaeicola_A_858004*, and *Copromonas* (phylum *Bacteroidota*) were negatively correlated with purine metabolites (e.g., adenosine, inosine, MTA, 1-methyladenosine), which were elevated in HF, indicating potential microbial regulation of host purine metabolism and methylation balance.

To account for potential confounding variables, we performed graph-based causal inference analysis, identifying 225 microbe–metabolite interaction pairs in the HF network—substantially more than the 91 pairs in controls (**Figure 4A, Table S14-15)**. Notably, the HF network had a higher proportion of negative regulatory interactions (41.3%, 93 out of 225 pairs) compared with the control group (19.8%, 18 out of 91 pairs). In particular, gut microbial taxa such as *Phocaeicola_A_858004*, *Clostridium_Q_135822*, *Enterocloster*, and *Alistipes_A_871404* exerted significant negative causal effects on key metabolites, including MTA, aceglutamide (Gln), 1-methyladenosine, phenylacetylglutamine, and kynurenine. Subsequently, bi-directional mediation analysis validated 256 significant microbe–metabolite–HF linkages, as an additional filter and reinforcement of the causal interactions (*P*_mediation_ < 0.01 and *P*_inverse_ mediation > 0.1, **Figure S7E, Table S16**), with six pairs consistently supported by both causal inference and mediation analysis (**Figure 4B, Table S17)**. For example, *Phocaeicola_A_858004* influenced HF via MTA and Gln (**Figure 4C)**, while *Clostridium_Q_135822* acted through N6-threonylcarbamoyladenosine. Other notable pairs included *Lachnospira* via N,N-dimethylguanosine, *Barnesiella* via sebacic acid, and *Enterocloster* via PAGln **(Figure S7A-D)**. Despite arising from distinct pathways, these metabolites converge on core processes such as energy metabolism, oxidative stress, inflammation, and cardiomyocyte function ^[21]^. Hence, these findings underscore robust microbe–metabolite associations that may critically shape host metabolism and drive HF pathogenesis.

**Figure 4.**
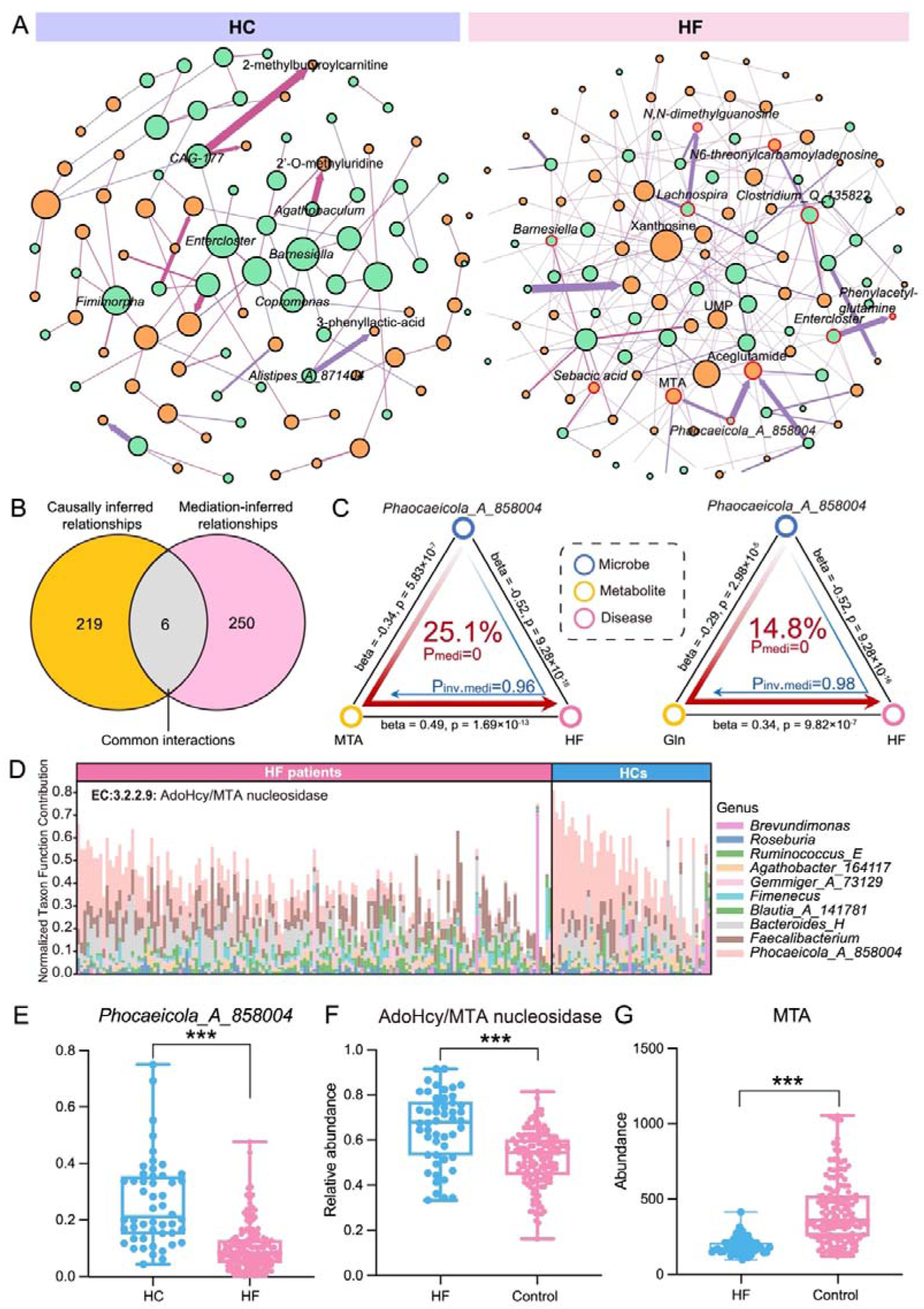
The causal crosstalk between gut microbiome and serum metabolites in HF patients and HCs. **(A)** Causal interaction networks in HF patients and HCs inferred by causal inference. Node colors represent feature classes: genera (green) and metabolites (yellow). Red and purple edges denote positive and negative interactions, respectively (|correlation| > 0.4, FDR < 0.05). Node size indicates degree, and edge width reflects correlation strength. **(B)** Venn diagram of relationships identified by causal inference and mediation analysis. **(C)** Causal pathway from genus *Phocaeicola_A_858004* to HF via serum MTA and Gln. **(D)** Differential abundance of metagenomically inferred enzyme EC:3.2.2.9 in HF patients, annotated with contributing genera. Stacked bars represent normalized taxon-specific contributions per sample (N = 199), sorted by dominant contributor and grouped by phenotype (HF vs. HC). **(E–G)** Abundances of *Phocaeicola_A_858004*, enzyme EC:3.2.2.9, and serum MTA in HF patients and HCs.

To uncover potential mechanistic links among gut microbes and serum metabolites, we cross-referenced differential enzymes from fecal 16S data with those enriched in serum metabolites, revealing 41 shared enzymes **(Figure S8A)**. Surprisingly, *Phocaeicola_A_858004* emerged as the primary contributor to 5’-nucleotidase and AdoHcy/MTA nucleosidase, key enzymes regulating nucleoside metabolism and linked to metabolites including MTA, guanosine, adenosine, and UMP **(Figure S8B)**. Notably, among all metabolites analyzed, only MTA was directly mapped to a microbial metabolic pathway in our functional analysis—the S-adenosyl-L-methionine (SAM) cycle I. MTA is a sulfur-containing nucleoside that is generated as a by-product during polyamine biosynthesis from SAM, a central methyl donor in cellular methylation reactions ^[22, 23^]. Within this pathway, the enzyme AdoHcy/MTA nucleosidase plays a pivotal role by catalyzing the hydrolysis of MTA, thereby regulating its degradation and preventing excessive accumulation. In HF patients, both *Phocaeicola_A_858004* abundance and its contributions to the SAM cycle I and AdoHcy/MTA nucleosidase were significantly reduced (**Figure 4C, 4E; Figure S8C–D)**, accompanied by elevated serum MTA levels (**Figure 4G)**. Correlation analyses further confirmed functional interdependence among *Phocaeicola_A_858004*, AdoHcy/MTA nucleosidase, and serum MTA **(Figure S8E–G)**. Together, these data indicate that reduced *Phocaeicola_A_858004* in HF diminishes microbial MTA degradation via AdoHcy/MTA nucleosidase, contributing to systemic MTA accumulation.

### *Phocaeicola vulgatus* reduces serum MTA levels and improves myocardial mitochondrial function

Within the genus *Phocaeicola_A_858004* in our data, *P. vulgatus* was the most abundant species (**Figure 5A)**. Given previous reports of reduced *P. vulgatus* abundance in coronary heart disease and HF patients ^[24, 25^], we focused on this species to validate the causal relationship among *Phocaeicola_A_858004*, MTA, and HF. Then, mouse models were established with three groups: sham-operated controls (Sham), HF model (HF-M), and HF mice receiving *P. vulgatus* gavage (HF-PV) (**Figure 5B)**. Consistent with human 16S data, HF-M mice exhibited a significant reduction in fecal *P. vulgatus*, whereas HF-PV mice showed markedly increased abundance, confirming successful colonization **(Figure S9A-B)**. Moreover, echocardiography revealed impaired cardiac function in HF-M mice, which was substantially improved by *P. vulgatus* administration, as indicated by higher left ventricular ejection fraction (LVEF%) and fractional shortening (LVFS%) (**Figure 5C–E)**. Histological analyses further demonstrated that *P. vulgatus* administration mitigated myocardial disorganization and inflammatory infiltration observed in HF-M mice, as evidenced by H&E and Masson’s staining, as well as reduced ANP/BNP expression (**Figure 5G, S9C-F)**. Considering the central role of energy supply dysbiosis in HF pathogenesis, we next examined myocardial mitochondrial integrity of mice in all groups. Specifically, transmission electron microscopy revealed swollen mitochondria with sparse, fragmented cristae and localized matrix dissolution in HF-M mice, whereas *P. vulgatus* colonization largely restored normal ultrastructure (**Figure 5H)**. Immunofluorescence further showed reduced ROS accumulation and preserved Tom20 expression in HF-PV hearts (**Figure 5I–L)**, demonstrating that *P. vulgatus* protects myocardial mitochondrial structure and function. Collectively, these results indicate that *P. vulgatus* administration improves cardiac function and attenuates myocardial mitochondrial dysfunction in HF mice.

**Figure 5.**
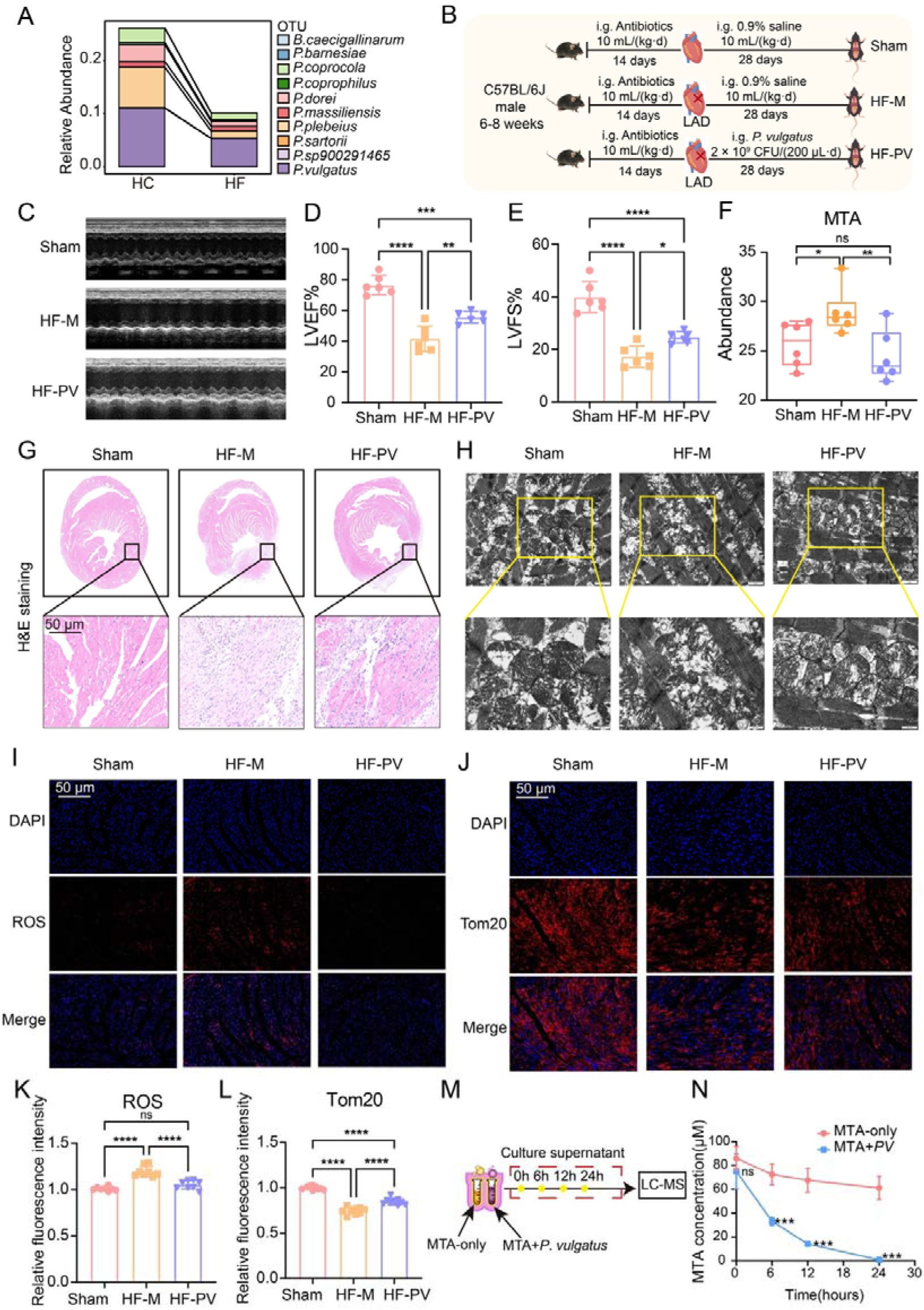
Validation of the effects of *P. vulgatus* on cardiac and mitochondrial function via modulation of MTA *in vivo* and *in vitro*. **(A)** Relative abundance of species within the genus *Phocaeicola_A_858004* in the gut microbiome of HF patients and HCs. **(B)** Experimental timeline for Sham, HF-M, and HF-PV mouse model construction and sample collection. **(C–E)** Representative echocardiographic images and quantification of LVEF% and LVFS% (n = 6). **(F)** Box plot showing serum MTA abundance in mice (n = 6). **(G)** Representative H&E staining of mouse myocardial tissue (n = 3). **(H)** Transmission electron micrographs illustrating mitochondrial ultrastructure in myocardial tissue. **(I-L)** Representative immunofluorescence images and quantification of ROS (I, K) and Tom20 (J, L) expression in myocardium, respectively. Fluorescence intensity was quantified as mean gray value per field and normalized to the Sham group (n = 3 hearts, 8 fields total). Scale bar = 50 μm. **(M)** Schematic workflow for LC–MS analysis of MTA content in bacterial culture supernatants. **(N)** Quantification of MTA levels in culture media by LC–MS (n = 3). Statistical significance was determined by one-way ANOVA for panels D, E, F, K, and L, and by two-way ANOVA for panel N. Significance levels are denoted as follows: *P* < 0.05 (*), *P* < 0.01 (**), *P* < 0.001 (***), *P* < 0.0001 (****), not significant (ns).

We then performed targeted metabolomics on serum samples of mice in all groups, identifying 22 differential metabolites in HF-M versus Sham mice, including AMP, L-lactic acid, 1-pentadecanoyl-sn-glycero-3-phosphocholine, and MTA, all linked to myocardial energy metabolism **(Table S18)**. KEGG enrichment highlighted phenylalanine metabolism as a key pathway **(Table S20, Figure S10A)**. In HF-PV mice, 32 metabolites differed relative to HF-M, including 11-dehydrocorticosterone, 3-methoxyindole, and 2-piperidone, associated with glutathione metabolism and arginine/proline metabolism **(Tables S19, Table S21, Figure S10B)**. Importantly, serum MTA was significantly elevated in HF-M mice but returned to Sham levels upon *P. vulgatus* gavage (**Figure 5F)**, suggesting that *P. vulgatus* alleviates pathological MTA accumulation in HF. However, this finding alone does not establish a direct biological interplay between *P. vulgatus* and MTA. To address this, we further cultured *P.*LJ*vulgatus* in MTA-supplemented medium and monitored MTA concentrations over time using LC-MS (**Figure 5M)**. Notably, MTA levels decreased progressively over 24 h in the presence of *P. vulgatus*, while remaining stable in control medium without bacteria without *P.*LJ*vulgatus* (**Figure 5N)**. These findings demonstrate that *P. vulgatus* could actively consumes MTA, and its reduction may therefore contribute to serum MTA accumulation in HF.

### MTA aggravates HF by inducing myocardial stress response and mitochondrial impairment

To investigate the direct role of MTA in HF progression, we performed both *in vivo* and *in vitro* experiments. Mice were divided into three groups: Sham, HF model (HF-M), and HF-M mice receiving intraperitoneal MTA (30 mg/kg; HF-MTA) (**Figure 6A)**. Specifically, echocardiography revealed that MTA further impaired cardiac function in HF-M mice (**Figure 6B–D)**, accompanied by upregulation of myocardial ANP and BNP expression **(Figure S11A–B)**. Histological analysis showed that MTA exacerbated inflammatory infiltration and disorganized cardiomyocyte architecture (**Figure 6E)**. Transmission electron microscopy revealed worsened mitochondrial swelling, cristae disruption, and vacuolation in HF-MTA mice (**Figure 6F)**, which was corroborated by immunofluorescence of Tom20 and ROS, indicating intensified mitochondrial dysfunction (**Figure 6G–J)**. *In vitro*, H9c2 cells were subjected to 8-hour OGD with or without 100 μM MTA (**Figure 6K)**. Consistent with *in vivo* findings, MTA increased ANP and BNP expression in H9c2 cells **(Figure S11C–D)**. JC-1 staining showed a decreased red/green fluorescence ratio under hypoxia, reflecting reduced mitochondrial membrane potential, which was further exacerbated by MTA treatment (**Figure 6L-M)**. Moreover, flow cytometry confirmed that MTA enhanced mitochondrial membrane potential depolarization under hypoxic conditions (**Figure 6N, Figure S11E–G)**.

**Figure 6.**
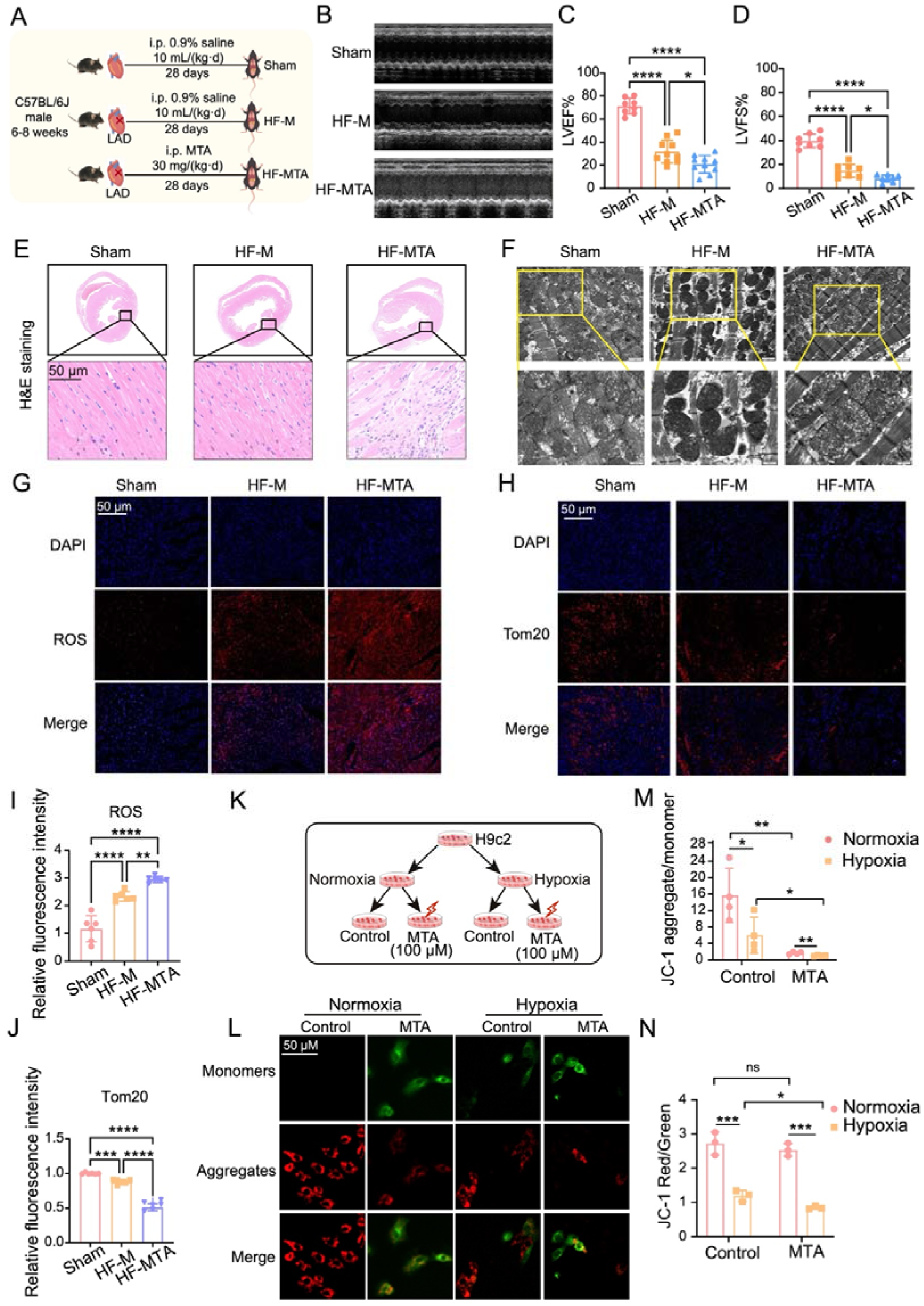
Assessment of cardiac and mitochondrial function under MTA treatment *in vivo* and *in vitro*. **(A)** Experimental timeline for Sham, HF-M, and HF-MTA mouse model construction and sample collection. **(B–D)** Representative echocardiographic images and quantification of LVEF% and LVFS% (n = 8). **(E)** Representative H&E staining of mouse myocardial tissue (n = 3). **(F)** Transmission electron micrographs illustrating mitochondrial ultrastructure in myocardial tissue. **(G-J)** Representative immunofluorescence images and quantification of ROS (G, I) and Tom20 (H, J) expression in myocardium, respectively. Fluorescence intensity was quantified as mean gray value per field and normalized to the Sham group (n = 3 hearts, 8 fields total). Scale bar = 50 μm. **(K)** Schematic illustration of the experimental design for treating H9c2 cells with vehicle or MTA (100 μM) under normoxic and hypoxic conditions. **(L-M)** Representative JC-1 staining images (L) and quantification of red/green fluorescence ratio (M) in H9c2 cells (n = 4). Red channel: JC-1 aggregates; green channel: JC-1 monomers; merge channel: overlay of red and green fluorescence. Scale bar = 50 μm. **(N)** Flow cytometric analysis of JC-1 red/green fluorescence ratio (n = 3). Statistical significance was determined by one-way ANOVA for panels C, D, I, and J, and by two-way ANOVA for panels M and N. Significance levels are denoted as follows: *P* < 0.05 (*), *P* < 0.01 (**), *P* < 0.001 (***), *P* < 0.0001 (****), not significant (ns).

Together, these results demonstrate that MTA promotes myocardial stress marker expression, aggravates inflammation and structural remodeling, and induces mitochondrial dysfunction both *in vivo* and *in vitro*. These findings support the hypothesis that reduction of *P. vulgatus* impairs myocardial mitochondria function by elevating systemic MTA, thereby contributing to the cardiac dysfunction in HF (**Figure 7)**.

**Figure 7.**
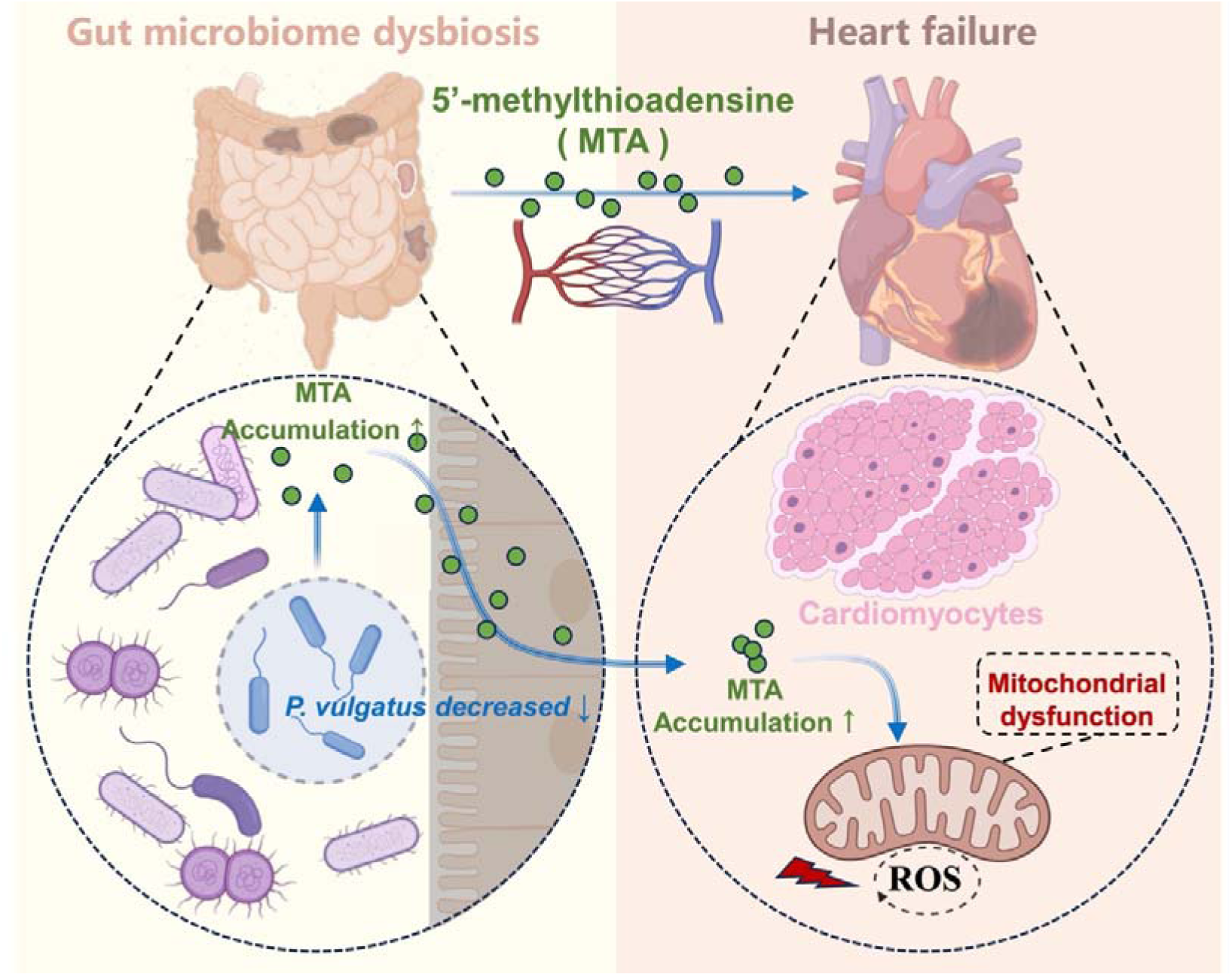
Depletion of *P. vulgatus* elevates circulating MTA, leading to mitochondrial dysfunction and HF progression. Schematic illustration of the gut–heart interplay. Reduction of *P. vulgatus* within the gut microbiota increases systemic MTA levels, which impairs mitochondrial oxidative phosphorylation in cardiomyocytes, resulting in energy metabolism disorders and promoting HF pathogenesis.

## Discussion

In this study, we systematically characterized the multidimensional alterations and diagnostic capabilities of gut microbiome and serum metabolome in HF patients in a Chinese cohort. Through causal inference, we identified key microbe–metabolite interactions with potential causal effects on HF progression, and further validated the major linkage experimentally *in vivo* and *in vitro*, thereby elucidating their pathogenic relevance in HF.

Consistent with the systemic metabolic dysregulation characteristic of HF, our integrated gut metagenomic and serum metabolomic analyses revealed widespread perturbations across microbial and host metabolic pathways. Microbial functions showed enhanced fermentation, nucleotide turnover, and fatty acid biosynthesis, paralleled by serum metabolomic enrichment in amino acid, nucleotide, lipid, and central carbon metabolism pathways. These concordant alterations indicate a coordinated disruption of energy homeostasis and redox balance along the gut–host axis. Importantly, both microbial and metabolic signatures demonstrated strong diagnostic potential for HF, surpassing conventional clinical biomarkers. Integrating multi-omics features with clinical parameters further improved diagnostic precision, reflecting the pivotal role of the gut and circulation as interconnected metabolic hubs in HF ^[5, 26, 27]^. Moreover, a simplified model retaining a minimal set of features achieved excellent performance (AUC = 0.91), underscoring the translational utility of integrative omics for precision diagnosis and mechanistic understanding of HF.

In exploring gut microbe–host metabolic interactions, we observed that the HF network exhibited enhanced microbe–metabolite connectivity, characterized predominantly by negative regulatory interactions involving nucleotide-related metabolites such as xanthosine, MTA, and UMP. These findings suggest a systemic remodeling of host–microbial metabolism in HF, marked by disrupted purine turnover, impaired methylation cycles, and altered microbial contributions. Causal inference and mediation analyses further identified *Phocaeicola_A_858004* as a potential upstream driver of HF, with MTA serving as a key metabolic intermediate linking microbial alterations to host metabolic dysfunction. In our cohort, HF patients displayed markedly reduced abundance of *Phocaeicola_A_858004* accompanied by elevated serum MTA levels (**Figure 4E, G)**, supporting this negative regulatory axis as a potential contributor to disease progression.

Among the taxa analyzed, *P. vulgatus* was identified as the dominant species within *Phocaeicola_A_858004*. As a core commensal in healthy gut ecosystems, *P. vulgatus* has been reported to exert protective effects in cardiovascular and metabolic diseases through modulation of inflammation and lipid metabolism ^[24, 28]^. Consistent with our previous findings ^[25]^, our results demonstrate that *P. vulgatus* supplementation markedly improved cardiac function, reduced pathological remodeling, and preserved myocardial mitochondrial structure and function in HF mice (**Figure 5)**. Notably, *P. vulgatus* administration reduced circulating MTA levels *in vivo*, while *in vitro* assays confirmed its capacity to directly consume MTA, firmly establishing its role in regulating host MTA homeostasis (**Figure 5F, M-N)**. Furthermore, we observed diminished contributions of *Phocaeicola_A_858004* to AdoHcy/MTA nucleosidase activity, implicating reduced microbial degradation of MTA in its systemic accumulation (**Figure 4D, F)**. This aligns with prior evidence showing that deletion of AdoHcy/MTA nucleosidase in E. coli causes intracellular MTA accumulation up to 50-fold higher than wild type ^[29]^. Collectively, these data suggest that depletion of *P. vulgatus* compromises microbial MTA catabolism, leading to accumulation of serum MTA in HF.

We further demonstrated that MTA aggravates HF progression by exacerbating inflammation and structural remodeling, as well as inducing mitochondrial dysfunction (**Figure 6)**. Consistent with our findings, previous studies have shown that MTA accumulation suppresses methyltransferase and polyamine synthase activities, thereby disrupting methylation balance, impairing cellular energy metabolism, and ultimately hindering cell growth—highlighting its detrimental effects when present at elevated levels ^[30, 31]^. In mammalian systems, excessive MTA has also been associated with disturbances in cAMP metabolism, endothelial adhesion molecule expression, and cytokine secretion ^[30]^. More importantly, high MTA levels contribute to mitochondrial dysfunction, characterized by impaired assembly of respiratory chain complexes, weakened antioxidant defenses, and reduced oxidative phosphorylation efficiency ^[32, 33]^. Thus, these findings identify MTA as a key mediator of mitochondrial impairment and a potential risk factor of HF progression. Taken together, we delineated a mechanistic pathway whereby depletion of *P. vulgatus* leads to the accumulation of serum MTA, which in turn induces mitochondrial dysfunction in cardiomyocytes and promotes HF progression, underscoring a potentially critical role of gut microbial regulation of adenosine-related metabolites in HF pathogenesis.

Moreover, causal analysis identified additional microbe–metabolite interactions converging on diverse yet HF-relevant biological processes. *Phocaeicola_A_858004* was also linked to glutamine (Gln), a key metabolic substrate essential for nucleotide and protein synthesis, vascular cell proliferation, and extracellular matrix remodeling. While adequate Gln availability protects against cardiometabolic stress, excessive glutaminolysis can promote maladaptive angiogenesis and vascular remodeling, thereby contributing to HF pathogenesis ^[34, 35]^. In parallel, *Enterocloster* was associated with PAGln—a well-characterized gut-derived metabolite that modulates adrenergic receptor signaling and is both clinically and mechanistically implicated in HF ^[36–38]^. Beyond these, *Clostridium_Q_135822* (via N6-threonylcarbamoyladenosine, tLA), *Lachnospira* (via N,N-dimethylguanosine), and *Barnesiella* (via sebacic acid) collectively highlight microbial influences on tRNA modification ^[39]^, nucleotide turnover ^[40]^, and fatty acid oxidation ^[41]^—processes closely linked to myocardial energy metabolism and HF development. Together, these findings expand the spectrum of potential microbe–metabolite axes involved in HF and provide valuable leads for future mechanistic and interventional studies.

Collectively, our integrative analysis reveals that gut microbial and metabolic signatures have superior diagnostic potential, facilitating earlier clinical detection for HF. This work further delineates the causal interplay between the gut microbiome and host metabolism, identifying key microbe–metabolite interactions that contribute to HF pathogenesis. Importantly, our findings underscore the value of integrating causal inference with experimental validation to elucidate pathogenic mechanisms and identify potential therapeutic targets for metabolic modulation in HF. Further multi-center longitudinal studies are warranted to validate our findings and to assess the translational potential of targeting the *P. vulgatus*–MTA axis for metabolic therapy in HF. Moreover, potential confounding from uncontrolled host intestinal genetic backgrounds needs to be addressed using family-based cohort ^[42]^.

## Conclusions

Our integrated multi-omics analysis delineates the multidimensional alterations and excellent diagnostic potential of the gut microbiome and serum metabolome in HF. Moreover, taking advantage of causal inference, we identify a novel mechanistic link between gut microbes and HF progression through the *P. vulgatus*–MTA axis, highlighting the therapeutic potential of targeting microbial metabolite pathways for clinical intervention.

## Materials and Methods

### Ethics approval and participant consent

All human study procedures were reviewed and approved by the Institutional Review Board at the Anzhen Hospital, Capital Medical University, Beijing (No. 2023142X). All participating individuals provided written informed consent prior to enrollment.

### Patient cohort and recruitment

A total of 199 age- and sex-matched Chinese participants were recruited from Anzhen Hospital, Capital Medical University, comprising 149 HF patients and 50 HCs. The detailed clinical characteristics of the cohort are summarized in **Table S1**.

Participants were aged ≥ 45 years and either had HF with prior myocardial infarction and/or angina (NYHA ≥ III and/or LVEF < 40%) or were healthy controls with normal ECG and no structural heart disease. All could complete questionnaires and provide fecal samples. Exclusion criteria included recent major gastrointestinal surgery, gastrointestinal diseases or infections (IBD, gastroenteritis, recurrent *C. difficile* or *H. pylori*), chronic GI conditions (diarrhea, constipation, ulcers, polyps, tumors, IBS, cholecystitis, hepatitis), recent use of antibiotics, probiotics, immunosuppressants, corticosteroids, or NSAIDs, or inability/unwillingness to participate.

### Sample collection and storage

Fecal and paired serum samples were collected from each participant. All samples were immediately frozen and stored at -80°C until DNA extraction and metabolomic analysis.

### 16S rRNA gene sequencing and microbiota profiling

Microbial genomic DNA was extracted from fecal samples using the cetyltrimethylammonium bromide (CTAB) method. The hypervariable V3-V4 region of the 16S rRNA gene was amplified with primers 341F (5’-CCTAYGGGRBGCASCAG-3’) and 806R (5’-GGACTACNNGGGTATCTAAT-3’). Amplified libraries were sequenced on an Illumina NovaSeq 6000 platform (Illumina, USA) by Wuhan Metware Biotechnology Co., Ltd.

Raw sequencing data were processed using the QIIME2 pipeline (version 2.7.1). Denoising, merging, and chimera removal were performed with DADA2 (via the q2-dada2 plugin) to generate amplicon sequence variants (ASVs). Taxonomic assignment was carried out using a naive Bayes classifier trained on the Greengenes2 database (2022.10 release).

### Microbial ecological and functional analysis

Alpha diversity (Shannon, ACE, Faith’s Phylogenetic, and Chao1 indices) and beta diversity (Bray-Curtis dissimilarity) were calculated using the R package vegan (v2.5.7). Differences in community structure were assessed by permutational multivariate analysis of variance (PERMANOVA) with 999 permutations. The functional potential of the microbial community was predicted from the 16S data using PICRUSt2 (Phylogenetic Investigation of Communities by Reconstruction of Unobserved States).

### Serum metabolomic profiling

Targeted metabolomics profiling of human and mouse serum samples was performed using the T500 Metabolomics Platform (Wuhan Metware Biotechnology Co., Ltd.). Metabolites were extracted with 20% acetonitrile–methanol, centrifuged, and filtered through a protein precipitation plate prior to analysis. Data acquisition was carried out on an ultra-performance liquid chromatography–tandem mass spectrometry (UPLC–MS/MS) system, and raw data were processed using Analyst (v1.6.3) and MultiQuant (v3.0.3) software for peak integration and quantification. Metabolite concentrations were calculated from standard curves and normalized for subsequent analysis using the R package MetaboAnalystR (v4.0).

For targeted quantification, LC–MS analysis was performed on a Waters Acquity UPLC I-Class/TQ-XS system equipped with an Acquity UPLC BEH C18 column (100 mm × 2.1 mm, 1.7 µm). The mobile phases consisted of 0.1% formic acid in water (A) and acetonitrile (B), with a flow rate of 0.3 mL·minL¹ and a column temperature of 35 °C. The gradient elution was set as follows: 5% B for 0.3 min, 5–90% B at 2.0 min, maintained for 0.5 min, and re-equilibrated to 5% B for 0.5 min. Electrospray ionization was operated in positive mode, and multiple reaction monitoring (MRM) was used for quantification.

### Differential signature analysis

To identify microbial taxa with significant differences in abundance between groups, we applied the LEfSe (Linear Discriminant Analysis Effect Size) method implemented in the R package “microeco” (v1.15.0), which integrates non-parametric statistical testing with linear discriminant analysis to determine both the statistical significance and biological relevance of differential signatures. Microbial signatures with LDA score > 2 and *P*-value < 0.05 were considered significantly different. For metabolic profiles, we employed Orthogonal Partial Least Squares Discriminant Analysis (OPLS-DA) within “MetaboAnalystR” (v4.0). Metabolites with VIP score > 1 and *P*-value < 0.05 were identified as significantly different between groups. Confounding effects were blocked by Wilcoxon test. For comparisons involving more than two groups, data were analyzed by one-way ANOVA. *P* < 0.05 was considered statistically significant.

### Multidimensional signatures association analysis

To ensure that microbial and metabolite associations were not confounded by demographic or clinical factors, permutational multivariate analysis of variance (PERMANOVA) was first performed to evaluate the effects of age, gender, BMI, and other clinical variables on feature abundance. Co-abundance patterns among differential microbial genera were then inferred using the SparCC algorithm ^[43]^, which estimates correlations from compositional data by simulating 500 Dirichlet-distributed iterations and computing pseudo *P-values* through bootstrap resampling. To further investigate cross-domain associations, Hierarchical All-against-All association testing (HALLA, v0.8.20) ^[44]^ was applied to identify significant correlations between microbial and metabolite signatures. Associations with an absolute correlation coefficient (|r|) > 0.4 and *P* < 0.05 were retained to construct the integrated microbe–metabolite interaction network, which was visualized using Gephi (v0.9.5).

### Diagnostic model construction and evaluation

A feedforward neural network (FNN) model was constructed using TensorFlow (v2.8.0) to discriminate HF patients from HCs based on multi-omics features. The model architecture included hidden layers with ReLU activation and an output layer with a sigmoid activation function. A dropout rate of 0.2 was applied to prevent overfitting. The model was trained using the Adam optimizer with a learning rate of 0.005.

Model performance was evaluated via stratified 10-fold cross-validation and on an independent testing set using metrics including the area under the receiver operating characteristic curve (AUC), accuracy, sensitivity, specificity, F1-score and normalized Matthews correlation coefficient (MCC) ^[45]^. Feature importance was interpreted using SHapley Additive exPlanations (SHAP) ^[46]^.

### Causal inference

Following an interventional causal inference algorithm based on Do-Calculus described previously ^[47, 48]^, we employed the Peter-Clark (PC) algorithm to infer the causal directed acyclic graph (DAG) among microbial genera and serum metabolites. The causal effect size for each directed edge in the network was quantified using the DoWhy library (v0.12) ^[49]^. Significance was assessed through permutation tests (999 permutations). Bidirectional mediation analysis (associational causal inference) was performed using the R package mediation (v4.5.0) to assess whether serum metabolites mediate the effect of gut microbes on HF status, controlling for age, sex, and BMI. The Benjamini-Hochberg procedure was used to control the false discovery rate (FDR).

### *P. vulgatus* culture and MTA utilization assay

*P. vulgatus* (ATCC 8482) was cultured anaerobically (10% COL, 5% HL, 85% NL) in Fluid Thioglycollate Medium. To assess MTA consumption, bacteria were inoculated into medium supplemented with 100 μM MTA. Supernatants were collected at 0, 6, 12, and 24 hours. MTA concentrations were quantified by LC-MS as described above.

### Mouse models of heart failure

All animal experiments were approved by the Animal Ethics Committee of Guangzhou University of Chinese Medicine (No. 20250509001) and conducted in compliance with the 3R principles.

#### HF model and bacterial supplementation

Male C57BL/6J mice (6–8 weeks old) were administered a broad-spectrum antibiotic cocktail (10 mL·kgL¹·dL¹, intragastrically) for 14 days to deplete gut microbiota. After antibiotic treatment, myocardial infarction–induced HF was established by permanent ligation of the left anterior descending (LAD) coronary artery. Sham-operated mice underwent the same procedure without LAD ligation. Following surgery, mice received daily intragastric administration for 28 days as follows: 0.9% saline (Sham and HF-M groups) or *P. vulgatus* suspension (2 × 10L CFU in 200 μL saline·dL¹, HF-PV group).

#### MTA administration study

In a separate experiment, mice received daily intraperitoneal injections for 28 days as follows: 0.9% saline (10 mL·kgL¹·dL¹, Sham and HF-M groups) or 5′-methylthioadenosine (MTA, 30 mg·kgL¹·dL¹, HF-MTA group).

### Echocardiography

On post-operative day 28, mice were anesthetized with isoflurane and transthoracic echocardiography was performed using a VINNO small-animal ultrasound system (Suzhou, China). Two-dimensional and M-mode images were acquired at the mid-papillary level in the parasternal short-axis view, left ventricular ejection fraction (LVEF%) and left ventricular fractional shortening (LVFS%) were measured.

### Histology and immunofluorescence

After euthanasia, mouse hearts were fixed in 4% paraformaldehyde, embedded in paraffin, and sectioned at 5 μm through the ligation site. Hematoxylin–eosin (H&E) and Masson’s trichrome staining were performed following standard protocols to evaluate myocardial morphology and fibrosis. For immunofluorescence, sections were deparaffinized, blocked with 10% goat serum, and incubated overnight at 4 °C with primary antibodies against TOM20 (1:400, A19403, Abclonal). After PBS washes, Cy3-conjugated secondary antibody (1:400, SeraCare) was applied for 1 h, and nuclei were counterstained with DAPI. ROS levels were assessed using a ROS detection kit (S0063, Beyotime) according to the manufacturer’s instructions. Images were acquired using a Leica fluorescence microscope, and fluorescence intensity was quantified with ImageJ.

### Transmission electron microscopy

Fresh myocardial tissue (∼1 mm³) was fixed in electron microscopy fixative (ASA1063, ASPEN) for 2–4 h, rinsed in PBS, dehydrated through graded ethanol, and embedded. Ultrathin sections were stained with uranyl acetate and lead citrate and examined under a transmission electron microscope (Tecnai G2 20 TWIN, FEI, USA).

### Cell culture and oxygen-glucose deprivation (OGD)

H9c2 cells were cultured in high-glucose DMEM supplemented with 10% FBS and 1% penicillin–streptomycin. Oxygen–glucose deprivation (OGD) was induced by incubating cells in serum- and glucose-free DMEM under hypoxic conditions (37 °C, 8 h). Mitochondrial membrane potential was measured using JC-1 dye (M8650, Solarbio) following the manufacturer’s protocol. Fluorescence was analyzed by flow cytometry (Agilent) and fluorescence microscopy (Olympus).

### Measurement of mitochondrial membrane potential

Mitochondrial membrane potential in cell was assessed using JC-1 (M8650, Solarbio, Beijing, China). Briefly, cells were detached with 0.25% trypsin-EDTA (3062782, Gibco, UK) pelleted at 800 rpm for 5 min at room temperature, and washed three times with PBS. The pellet was resuspended in 0.5 mL JC-1 solution and incubated for 20 min at 37 °C in the dark. After three washes with 1 mL JC-1 buffer, stained cells were analyzed with flow cytometry (Agilent, USA) and fluorescence microscope (Olympus, Japan).

### Quantitative real-time PCR (qRT-PCR)

Total RNA was extracted from heart tissues or cells using FreeZol Reagent (R711, Vazyme, Nanjing, China). cDNA was synthesized and amplified using BrightCycle Universal SYBR Green qPCR Mix (Abclonal) on a QuantStudio system (Thermo Fisher Scientific). Gene expression was normalized to GAPDH and calculated using the 2^(-ΔΔCt) method. Primer sequences are listed in **Table S22**.

### Statistical analysis

Unless otherwise specified, group comparisons for continuous variables were performed using the Wilcoxon rank-sum test (for two groups) or one-way ANOVA (for multiple groups). Categorical data were compared using the Chi-squared test. A two-sided *P-value* < 0.05 was considered statistically significant. All statistical analyses were performed in R (v4.1.0 or above).

## Data Availability

All data produced in the present study are available upon reasonable request to the authors

## Authors’ contributions

CL, WW and YW conceived and designed the project. SG performed the multi-omics analysis, AI modeling, and bioinformatics analysis. YL and ZD recruited the participants, collected the fecal and serum samples. MY, XS, YQ, HW, TC and ZY performed the experiments *in vivo* and *in vitro*. SG and MY drafted the manuscript. YL, YO, YW, WW, CL revised the manuscript. All authors read and approved the final manuscript.

## Data and code availability statement

All of the raw data in this study has been uploaded in the National Omics Data Encyclopedia under accession No. OEP00006636 and are available upon reasonable request. The data relevant to the study are included in the article or uploaded as supplementary information. The code and scripts are available on GitHub (https://github.com/github-gs/GutMicrobiome_SerumMetabolome_in_HF).

## Acknowledgements

This work was supported by the National Natural Science Foundation of China (82505223 to SG, 82230126 and U24A20800 to WW); the China Postdoctoral Science Foundation (2025M773866 to SG); National Key Research and Development Program of China (2022YFC3500100 to WW); National Science Fund for Excellent Young Scholars (82222075 to CL); Scientific Research Project of the Guangdong Provincial Administration of Traditional Chinese Medicine (20261089 to XS); the Beijing Natural Science Foundation (7252220 to YL); National TCM Heritage and Innovation Team Project (ZYYCXTD-C-202201); Guangdong Provincial Key Laboratory of Syndrome and Formula (2022B1212010012). The funders had no role in study design, data collection and analysis, decision to publish, or preparation of the manuscript.

## Abbreviations

AI: artificial intelligence
ANP: A-type natriuretic peptide
ASVs: exact sequence variants
BNP: B-type natriuretic peptide
CEV: Causal estimate values
ESI: Electrospray ionisation (ESI)
FDR: False discovery rate
FNN: feedforward neural network
Gln: aceglutamide
HALLA: Hierarchical All-against-All association testing
HC: Healthy control
H&E: Hematoxylin and eosin stain
HF: Heart failure
hsTnI: High-sensitivity troponin I
KO: KEGG Orthology
LVEF: Left ventricular ejection fraction
LVFS: Left ventricular fractional shortening
MCC: Matthews correlation coefficient
MTA: 5’-methylthioadenosine
MRM: Multiple Reaction Monitoring
OGD: Oxygen–glucose deprivation
ORA: Over-representation analysis
OPLS-DA: Orthogonal Partial Least Squares Discriminant Analysis
PAGln: Phenylacetylglutamine
PCoA: Principal coordinate analysis
RFE: recursive feature elimination
PERMANOVA: permutational multivariate analysis of variance
ReLU: rectified linear unit
SAM: S-adenosyl-L-methionine
SHAP: SHapley Additive exPlanations
t6A: N6-threonylcarbamoyladenosine
PC: Peter-Clark
UPLC-MS/MS: Ultra-performance liquid chromatography coupled to tandem mass spectrometry

